# Testing the Validity of the Modified Vaccine Attitude Question Battery across 22 Languages with a Large-scale International Survey Dataset: Within the Context of COVID-19 Vaccination

**DOI:** 10.1101/2021.12.06.21267289

**Authors:** Hyemin Han

## Abstract

In this study, we tested the validity of the modified version of the Vaccine Attitude Question Battery (VAQB) across 22 different languages. Validity test was conducted with a large-scale international survey dataset, COVIDiSTRESSII Global Survey, collected from 20,601 participants from 62 countries. We employed exploratory and confirmatory factor analysis, measurement invariance test, and measurement alignment for internal validity test. Moreover, we examined correlation between the VAQB score, vaccination intent, compliance with preventive measures, and trust in public health-related agents. The results reported that the modified VAQB, which included five items, showed good validity across 22 languages with measurement alignment. Furthermore, the VAQB score showed negative association with vaccination intent, compliance, and trust as expected. The findings from this study provide additional evidence supporting the validity of the modified VAQB in 22 languages for future large-scale international research on COVID-19 and vaccination.

## Introduction

COVID-19 vaccines have been reported as an effective and reliable mean to prevent COVID-19 caused hospitalization and death across the globe ^1^. Although scientific research has consistently supported their safety and effectiveness, negative attitudes toward vaccines have become a serious issue ^2^. Because how to deal with such negative attitudes is one of the most fundamental problems that shall be addressed to promote wide distribution of the COVID-19 vaccines, and eventually, to end the COVID-19 pandemic, it is necessary to have a reliable and valid way to examine vaccine attitude. For example, ^3^ developed and tested the Vaccination Hesitancy Scale (VHS) consisting of ten items; in the study, the VHS was validated in two languages, English and French, among Canadian populations. Since its development, the VHS has been widely employed in studies focusing on people’s attitude on vaccination ^4^.

Although the VHS has been tested in validated in prior research, whether it can be applicable across different languages in a consistent way has not been fully examined. Given the COVID-19 pandemic is a global issue, international and cross-cultural research is strongly required. The majority of the previous studies employing the VHS tested the scale within a single-language study context, so they would not sufficiently support cross-language validity of the scale, which requires for international and cross-cultural comparisons or investigations ^5^. Similarly, other scales that have been less employed than the VHS have not been well tested for their validity in multi-lingual settings (e.g., ^2^).

In this study, we aimed at testing the validity of the Vaccine Attitude Question Battery (VAQB), which was modified from the original VAQB employed in a large-scale survey regarding vaccine attitude conducted in 2020 ^2^, across different languages by examining a large-scale dataset, COVIDiSTRESSII Global Survey (COVIDiSTRESSII), collected 20,601 participants from 62 countries ^6^. First, we performed exploratory and confirmatory factor analysis (EFA and CFA) with the English version of the modified VAQB. Second, we conducted measurement invariance (MI) test across 22 languages to examine whether the measurement structure of the modified VAQB was valid across different languages. Third, we performed measurement alignment to address the existing measurement non-invariance. Finally, we examined correlation between the calculated VAQB score and other variables that are expected to be positively associated with positive vaccine attitude, i.e., vaccination intent, compliance with preventive measures, trust in public health-related agents ^7,8^.

## Methods

### Analyzed Dataset

COVIDiSTRESSII dataset was collected from 20,601 participants using 48 different languages across the globe (see Supplementary Methods for further details about translation, data collection, and data preprocessing procedures) ^6^. For MI test and measurement alignment, which involve CFA, only the language versions with *N* ≥ 200 were analyzed. As a result, responses from 14,271 participants using 22 different languages were used in the present study (see Table S1 for the list of languages and demographics). Further details about the original dataset collection and preprocessing procedures are available in COVIDiSTRESSII project page, https://osf.io/36tsd. COVIDiSTRESSII project was reviewed and approved by the Research, Enterprise and Engagement Ethical Approval Panel at University of Salford (ref.: 1632).

### Measures

All measures were translated into different languages by COVIDiSTRESSII Consortium members. Further details about the employed measures are available in Supplementary Methods.

#### Modified Vaccine Attitude Question Battery

The modified VAQB with six item was employed to assess participants’ attitude to get COVID-19 vaccines. The six items were extracted and modified from the original VAQB ^2^ for a feasibility within the context of the large-scale international survey project based on discussions between COVIDiSTRESSII Consortium members. Responses were anchored to a seven-point Likert scale (1—strongly disagree to 7— strongly agree). Four items, Items 4 and 5 were reverse coded.

#### Vaccination Intent

Intent to get COVID-19 vaccines was assessed one item. Responses were anchored to a five-point Likert scale (1—not willing at all to 5—very willing).

#### Items for Compliance with Preventive Measures

We surveyed participants’ compliance with non-pharmaceutical measures to prevent spread of COVID-19. These items were adapted from the previous round of COVIDiSTRESS Global Survey ^9,10^. We examined compliance in three different behavioral domains, i.e., indoor and outdoor mask use, social distancing. Each type of compliance was measured with one item. Responses were anchored to a seven-point Likert scale (1—strongly disagree to 7—strongly agree).

#### Items for Trust

Participants’ trust in four different agents, i.e., health system, the World Health Organisation (WHO), governmental effort to handle the pandemic, science research, related to vaccination was also assessed. Similar to the items for compliance, the trust items were also adapted from COVIDiSTRESS Global Survey ^9,10^. Trust in each agent was surveyed with one item. Responses were anchored to a 11-point Likert scale (0—no trust to 10—complete trust).

### Analysis Plan

In this section, we described how the reliability and validity of the modified VAQB were tested. All R source code and data files are available to the public via the Open Science Framework, https://doi.org/10.17605/OSF.IO/QCPZX.

#### Exploratory and Confirmatory Factor Analysis

The whole English version dataset was randomly separated into two subsets, one for EFA (50%) and one for CFA (50%), to prevent overfitting. Then, we performed EFA of the modified VAQB with the first subset with R packages, *EFA*.*dimensions* and *EFAtools*. First, we tested whether EFA can be adequately performed with Kaiser-Meyer-Olkin (KMO) and Bartlett’s test. Second, we employed diverse measures, i.e., parallel analysis (PA), minimum average partial (MAP) test, hull method, Kaiser-Guttman criterion (KGC), to determine the number of factors to be explored ^11^. Third, we performed EFA with the determined factor number. In general, we assumed that factor loadings smaller than .50 as inappropriate ^12^.

Then, we conducted CFA with the second subset. CFA was performed with the measurement model identified by EFA with *lavaan* package. Because responses were anchored to a Likert scale, we employed *WLSMV* estimator, which is suitable for CFA with ordinal responses. CFA was conducted again with the whole data with the model. I examined whether RMSEA and SRMR < .08 and CFI and TLI ≥ .90 at the least ^13^. Furthermore, we also investigated whether each factor loading was significant at *p* < .05 ^12^. If the aforementioned requirements for EFA and CFA indicators were not fulfilled, we revised the modified VAQB by adjusting items and examined the fit indicators once again with the modified version.

Additionally, we examined the internal consistency of the modified VAQB across different languages in term of Cronbach α. The brief descriptive statistics and internal consistency of the modified VAQB in α for each language version are presented in Table S1.

#### Measurement Invariance Test

MI test was performed to examine whether the measurement model of the modified VAQB validated in the English version can also be validated across different language versions. It was performed by setting the group variable and equal parameter conditions in *lavaan*.

There are different levels of MI depending on the different equal parameter assumption. First, configural invariance only assumes the equal measurement structure across different groups. Second, metric invariance additionally assumes that factor loadings are equal. Third, scalar invariance additionally requires equal intercepts. Finally, the strictest invariance, scalar invariance, is achieved when the equal residual assumption is additionally satisfied ^14^.

Whether a specific level of invariance was achieved was examined by comparing fit indicators, i.e., RMSEA, SRMR, CFI, TLI, between two different levels of invariance. In the case of metric invariance, indicator changes should be smaller than -.01 CFI, +.015 RMSEA, and +.30 SRMR. For scalar invariance, changes should be less than -.01 CFI, +.015 RMSEA, and +.15 SRMR ^15^. For between-group comparison, scalar invariance must be satisfied at the least ^5^.

#### Measurement Alignment

If scalar invariance was not supported in the MI test, we performed measurement alignment to address the existing non-invariance. We employed *sirt* package to implement measurement alignment in R ^5^. Measurement alignment is a procedure to adjust factor loadings, intercepts, and group means to address non-invariance ^5,16^. To examine whether non-invariance was successfully addressed, we checked whether the resultant *R*^2^_loadings_ and *R*^2^_intercepts_, which indicate to what extent the non-invariance was absorbed via measurement alignment, approached 100% ^16,17^. If both *R*^2^ values exceeded 95% as shown in the prior research using measurement alignment ^18^, we assumed achievement of scalar invariance through measurement alignment ^16,17^.

Then, we calculated an adjusted VAQB factor score for each language group with adjusted factor loadings and intercepts. The factor score was calculated with the pseudo inverse matrix of factor loadings with *MASS* package ^19^.

#### Correlation Analysis

We conducted correlation analysis to acquire additional evidence supporting the convergent validity of the modified VAQB. Correlation between the VAQB score and other surveyed variables, vaccination intent, compliance, and trust, which were supposed to be positively associated with vaccine attitude ^7,8^, was examined. We assumed that the convergent validity of the VAQB was supported once significant positive correlation between the vaccine attitude, vaccination intent, compliance, and trust was found. For additional information, correlation between the VAQB items was also investigated.

## Results

### Exploratory and Confirmatory Factor Analysis

Both KMO, .82, and Bartlett’s test, χ^2^(15) = 1,545.28, *p* < .001, indicated that EFA was able to be adequately performed with the current scale and first English-version subset. All factor number determination methods, i.e., PA, MAP test, hull method, KGC, unequivocally suggested that only one factor was sufficient in the measurement model. The result of EFA with the one-factor model is presented in Table 1. Because the factor loading of Item 4 did not exceed the cutoff, .50, we conducted CFA with caution.

**Table 1.** Standardized factor loading resulting from EFA and CFA

| 6-item VAQB | 5-item VAQB |
| --- | --- |

|  | EFA<br>(English) | CFA<br>(English) | CFA<br>(English) | CFA<br>(whole) |
| --- | --- | --- | --- | --- |
| Item 1 | .78 | .79 | .84 | .89 |
| Item 2 | .70 | .76 | .79 | .81 |
| Item 3 | .58 | .60 | .63 | .60 |
| Item 4 | .44 | .42 | - | - |
| Item 5 | .61 | .59 | .51 | .58 |
| Item 6 | .78 | .64 | .66 | .84 |

When CFA was performed with the second English-version subset, the original measurement model including all six items did not report good model fit indicators, RMSEA = .15, PCLOSE = .00, SRMR = .06, CFI = .75, TLI = .58. Thus, we excluded the fourth item that showed the lowest standardized factor loading (.42) and did not show the satisfactory factor loading from EFA.

Hence, we performed CFA once again with the updated version with five items. The modified five-item scale well fitted the data, RMSEA = .04, PCLOSE = .69, SRMR = .02, CFI = .99, TLI = .98. All factor loading reported *p* < .001. When CFA was performed with the whole dataset with the five-item model, acceptable model fit indicators were reported as well, RMSEA = .07, PCLOSE = .00, SRMR = .02, CFI = .98, TLI = .96. Similar to the case of the English version, all factor loadings demonstrated *p* < .001. All resulting standardized factors loadings are presented in Table 1.

### Measurement Invariance Test

When configural invariance was tested, the resultant model fit indicators suggested mediocre fit, RMSEA = .09, PCLOSE = .00, SRMR = .03, CFI = .93, TLI = .87. With the equal loading assumption, metric invariance was tested. However, the model did not fit data well, RMSEA = .11, PCLOSE = .00, SRMR = .09, CFI = .81, TLI = .78. Additionally, the large changes in RMSEA (+.02), SRMR (+.06), CFI (-.12), and TLI (-.08), suggested metric invariance could not be achieved. Given scalar invariance, which is minimally required for between-group comparison, could not be supported by data, we performed measurement alignment to address the non-invariance across different languages.

### Measurement Alignment

The results from measurement alignment demonstrated that measurement non-invariance across different languages was successfully resolved by adjusting factor loadings and intercepts. The resultant *R*^2^_loadings_ = .97 and *R*^2^_intercepts_ = .98. They indicate that approximately 100% of existing non-invariance was absorbed by latent factor means and variances varying across languages, and thus, scalar invariance can be achieved via measurement alignment.

### Correlation Analysis

Correlation between surveyed variables, the VAQB factor score, general vaccination intent, compliance, and trust, is presented in Table 2. As expected, vaccine attitude was positively associated with all other variables, so the convergent validity of the modified VAQB was supported. Moreover, all VAQB items showed significant correlation with each other (see Table S2).

**Table 2.**
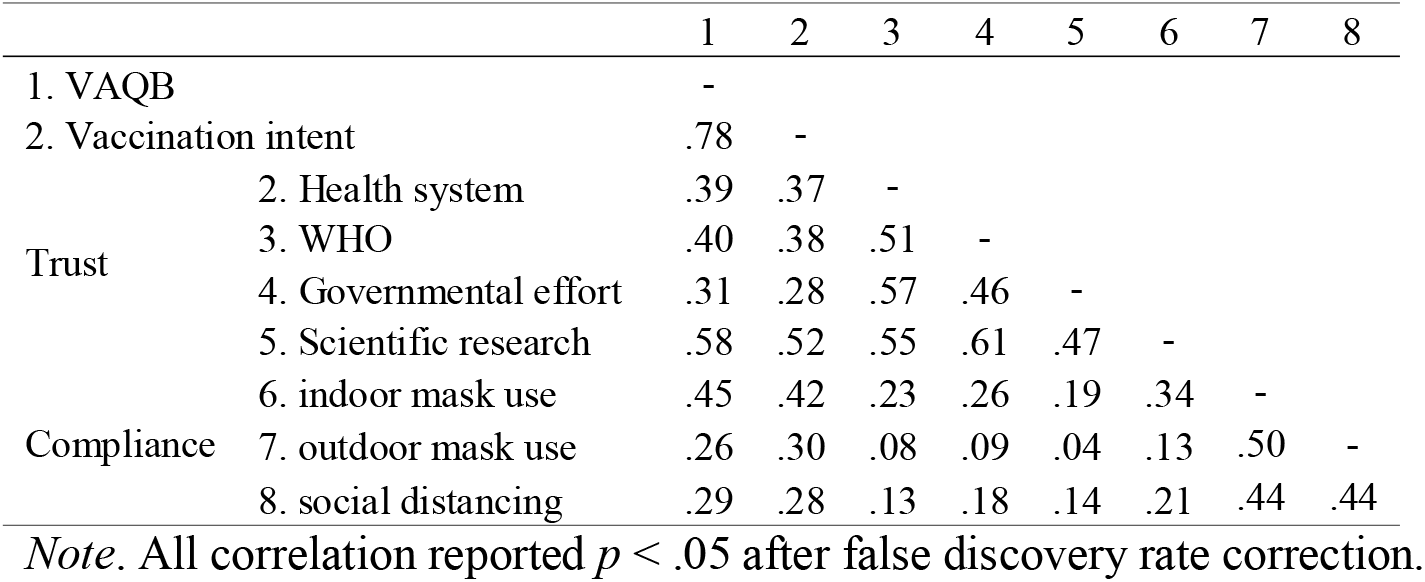
Correlation between the VAQB factor score and indicators related to vaccination intent, trust, and compliance with non-pharmaceutical preventive measures

## Discussion

In this study, we tested the validity of the modified VAQB across 22 different languages. We found that the original measurement model should be modified to achieve good model fit, so one item was excluded from the original six-items scale. The five-item scale reported acceptable model fit. However, measurement non-invariance was reported from the MI test. When measurement alignment was performed, the existing non-invariance was successfully absorbed; the large resultant *R*^2^ values suggest achievement of scalar invariance through measurement alignment ^18^. The correlation analysis demonstrated that the calculated factor score of vaccine attitude after alignment was positively associated with general vaccination intent, compliance with non-pharmaceutical preventive measures, and trust in health organizations, governmental efforts, and science research, in line with previous research ^7,8^.

Although the findings suggested the potential utility of the modified VAQB, several limitations warrant further investigations. First, although our dataset is a large-scale international survey dataset, it has been collected via convenient samples (i.e., internet users). Thus, the generalizability of the findings could be limited due to the sample quality and bias issues. Second, we employed self-report measures for vaccine intent and compliance, so predictive validity in terms of whether the measured vaccine attitude can predict actual behavioral outcomes could not be fully supported.

## Conclusion

The modified VAQB, which contains five items, can be reliably and validly administrated in 22 different languages with assistance of measurement alignment addressing measurement non-invariance. Because the version with six items did not show good psychometrical quality, we excluded one item and re-tested the version with five items. The correlational analysis also provides additional evidence supporting the convergent validity of the scale. In conclusion, the modified VAQB will be able to be widely utilized in future large-scale international studies regarding COVID-19 vaccination, particularly those involving cross-cultural or cross-national comparisons, with measurement alignment.

## Data Availability

All R source code and data files are available to the public via the Open Science Framework, https://doi.org/10.17605/OSF.IO/QCPZX.

## Acknowledgements

The author thanks Sara Vestergren and COVIDiSTRESSII Global Survey Consortium members for their help regarding data collection and preprocessing. Disclosure statement: No potential conflict of interest was reported by the author.

