## Supplementary Methods and Tables for "Testing the Validity of the Modified Vaccine Attitude Question Battery across 22 Languages with a Large-scale International Survey Dataset: Within the Context of COVID-19 Vaccination"

**COVIDiSTRESSII Global Survey Dataset**

The COVIDiSTRESSII Global Survey Dataset was collected by the COVIDiSTRESSII Global Survey consortium members consisting of experts in psychology, public health, political science, economics, and COVID-19-related social sciences across the globe. For international administration of the survey, all items in the survey form were translated and back translated by the consortium members. The English version of each scale was translated and then back translated for verification by 150 international scholars who speak 48 languages as native speakers (for the full list of the languages and further details about the consortium and translation process, refer to <https://osf.io/428pz/>).

Participants were recruited across the globe via the internet (e.g., Facebook, Twitter, Instagram, local internet bulletin boards, etc.) by the consortium members. Once they agree to participate in the survey, they were provided with a Qualtrics link. Then the translated survey forms were presented to the participants via Qualtrics. The collected data was preprocessed through the data cleaning procedures described in <https://osf.io/428pz/>. The data cleaning procedures were performed to filter out responses without consent, test responses, responses completed via a preview link, responses from participants who failed the attention check, and very quick responses (< 3 minutes to complete). Consequently, responses collected from 20,601 participants were finally included in the cleaned dataset.

**Modified Vaccine Attitude Question Battery**

The modified Vaccination Attitude Question Battery (VAQB) was used to measure participants’ hesitation to get COVID-19 vaccines. The six items were extracted and modified from the original VAQB using ten items. The COVIDiSTRESSII Consortium members determined and revised the six items to be included in the survey project to improve the brevity and feasibility of the scale within the context of the large-scale international survey project.

First, participants were presented with a general direction:

*Now we want to ask you a few questions about your thoughts on the COVID-19 vaccine.*

*Please indicate for the following statements to what extent you agree with them.*

Then, they were presented with the six items:

1. *Getting vaccines is a good way to protect children from disease*
2. *Generally, I do what my doctor recommends about vaccines*
3. *New vaccines are recommended only if they are safe*
4. *I am concerned about serious side effects of vaccines* **(reverse coded)**
5. *Parents should have the right to refuse vaccines required for schools for any reason* **(reverse coded)**
6. *Vaccinations are one of the most significant achievements in improving public health*

**Vaccination Intent**

Intent to get COVID-19 vaccines was measured with one item. At the beginning, participants were presented with a general direction, “Now we want to ask you a few questions about your thoughts on the COVID-19 vaccine.” Then, they were presented with the one item, “How willing are you to get the vaccine if one becomes available to you?” Responses were anchored to a five-point Likert scale (1—not willing at all to 5—very willing).

**Items for Compliance with Preventive Measures**

Compliance with three different types of non-pharmaceutical preventive measures was also measured. One item was used for each behavioral domain. First, participants were presented with a general direction:

*Many countries have issued guidelines for staying safe during the COVID-19 pandemic. Think about the last month, to what extent did you:*

Then, they were presented with the three items:

1. *Wear a face covering in public when indoors (e.g. in a supermarket or cafe)* 🡪 indoor mask use
2. *Wear a face covering in public when outdoors (e.g. in the street or park)* 🡪 outdoor mask use
3. *Stay at least the recommended distance (for example 2 meters/6 feet) from* people who are not part of your household 🡪 social distancing

Responses were anchored to a seven-point Likert scale (1—strongly disagree to 7—strongly agree).

**Items for Trust**

Participants’ trust in four different agents related to public health was also measured. Trust in each agent was measured by one item. First, participants were presented with a general direction:

*Please tell us how much you trust each of the institutions below. Please base your answer on your general impression.*

Then, they were presented with the four items:

1. *[residing_country's] Health system?* 🡪 trust in health system
2. *The World Health Organisation (WHO)*  🡪 trust in the WHO
3. *[residing_country's] Government’s effort to handle Coronavirus*  🡪 trust in governmental efforts.
4. Scientific research community 🡪 trust in science research

Then, responses were anchored to a 11-point Likert scale (0—no trust to 10—complete trust).

**Supplementary Tables**

| **Table S1** |  |  |  |  |  |  |  |  |  |
| --- | --- | --- | --- | --- | --- | --- | --- | --- | --- |
| Demographics, and internal consistency and descriptive statistics of the Vaccine Attitude Question Battery | | | | | | | | | |
|  | N | Gender (%) | | | Age | | Vaccine Attitude Question Battery factor score with alignment | | |
|  |  | Female | Male | Other/ Would rather not say | Mean | SD | α | Mean | SD |
| Whole data | 14,271 | 67.13% | 31.86% | .10% | 36.59 | 14.45 | .86 | -.19 | 1.60 |
| English | 1,590 | 65.89% | 32.91% | 1.20% | 30.39 | 11.38 | .81 | .43 | 1.13 |
| Bulgarian | 291 | 74.57% | 25.09% | .34% | 41.77 | 16.47 | .83 | .00 | 1.78 |
| Czech | 374 | 71.39% | 26.74% | 1.87% | 33.89 | 11.35 | .87 | -.61 | 1.68 |
| German | 703 | 64.15% | 35.28% | .57% | 43.83 | 18.27 | .88 | .20 | 1.29 |
| Spanish (Colombia) | 572 | 67.66% | 31.82% | .52% | 39.81 | 12.43 | .80 | .78 | .96 |
| Spanish (Costa Rica) | 230 | 70.43% | 28.26% | 1.30% | 36.33 | 10.42 | .80 | .63 | .98 |
| Spanish (Ecuador) | 305 | 65.25% | 33.77% | .98% | 31.92 | 10.62 | .81 | .61 | 1.04 |
| Spanish (Spain) | 706 | 65.82% | 33.76% | .43% | 40.20 | 13.48 | .84 | .69 | 1.06 |
| Spanish (Guatemala) | 240 | 83.33% | 16.67% | .00% | 36.08 | 14.34 | .79 | .51 | 1.07 |
| Spanish (Uruguay) | 291 | 87.59% | 12.41% | .00% | 42.25 | 12.87 | .82 | .48 | 1.08 |
| Spanish (Honduras) | 435 | 66.67% | 32.41% | .92% | 25.83 | 8.17 | .77 | .35 | 1.05 |
| Estonian | 262 | 87.36% | 12.64% | .00% | 39.37 | 10.33 | .87 | .36 | 1.27 |
| Finnish | 921 | 78.61% | 19.87% | 1.52% | 46.22 | 14.45 | .89 | .85 | 1.31 |
| Italian | 320 | 73.98% | 25.71% | .31% | 44.52 | 16.17 | .86 | .20 | 1.46 |
| Japanese | 2,136 | 41.85% | 56.88% | 1.26% | 45.51 | 11.11 | .70 | -.46 | 1.05 |
| Norwegian | 373 | 81.77% | 17.96% | .27% | 40.29 | 13.20 | .88 | .61 | 1.22 |
| Portuguese (Portugal) | 485 | 70.93% | 27.84% | 1.24% | 33.55 | 15.01 | .80 | .52 | .83 |
| Portuguese (Brazil) | 468 | 72.01% | 27.56% | .43% | 38.27 | 13.15 | .65 | .20 | .33 |
| Russian | 2,747 | 71.63% | 26.95% | 1.42% | 27.24 | 11.39 | .79 | -2.09 | 1.54 |
| Slovak | 313 | 88.82% | 11.18% | .00% | 34.64 | 13.55 | .90 | .05 | 1.45 |
| Ukrainian | 241 | 64.32% | 34.85% | .83% | 32.10 | 10.26 | .84 | .64 | 1.15 |
| Chinese | 268 | 63.43% | 34.70% | 1.87% | 35.28 | 10.05 | .58 | .57 | .83 |

**Table S2**

Correlation between the VAQB items

|  | Item1 | Item 2 | Item 3 | Item 5 |
| --- | --- | --- | --- | --- |
| Item 2 | .73 |  |  |  |
| Item 3 | .52 | .54 |  |  |
| Item 5 | .54 | .46 | .31 |  |
| Item 6 | .73 | .66 | .52 | .51 |

*Note*. All correlation reported *p* < .001 after false discovery rate correction.
